# Application of ConvNeXt with Transfer Learning and Data Augmentation for Malaria Parasite Detection in Resource-Limited Settings Using Microscopic Images

**DOI:** 10.1101/2024.10.31.24316549

**Authors:** Outlwile Pako Mmileng, Albert Whata, Micheal Olusanya, Siyabonga Mhlongo

## Abstract

Malaria is one of the most widespread and deadly diseases across the globe, especially in sub-Saharan Africa and other parts of the developing world. This is primarily because of incorrect or late diagnosis. Existing diagnostic techniques mainly depend on the microscopic identification of parasites in the blood smear stained with special dyes, which have drawbacks such as being time-consuming, depending on skilled personnel and being vulnerable to errors.

This work seeks to overcome these challenges by proposing a deep learning-based solution in the ConvNeXt architecture incorporating transfer learning and data augmentation to automate malaria parasite identification in thin blood smear images. This study’s dataset was a set of blood smear images of equal numbers of parasitised and uninfected samples drawn from a public database of malaria patients in Bangladesh. To detect malaria in the given dataset of parasitised and uninfected blood smears, the ConvNeXt models were fine-tuned. To improve the effectiveness of these models, a vast number of data augmentation strategies was used so that the models could work well in various image capture conditions and perform well even in environments with limited resources. The ConvNeXt Tiny model performed better, particularly the re-tuned version, than other models, such as Swin Tiny, ResNet18, and ResNet50, with an accuracy of 95%. On the other hand, the re-modified version of the ConvNeXt V2 Tiny model reached 98% accuracy. These findings show the potential to implement ConvNeXt-based systems in regions with scarce healthcare facilities for effective and affordable malaria diagnosis.

## 1. Introduction

According to the World Health Organization (WHO), there were 241 million cases and 627,000 deaths due to malaria in 2020, with the disease primarily affecting low-income countries (1). Malaria is an infectious disease caused by parasitic protozoa of the genus *plasmodium*, of which *plasmodium falciparum* and *plasmodium vivax* are the most pathogenic to man (2). These are transmitted through the bites of infected *Anopheles* mosquitoes. In malaria-endemic areas, timely and correct diagnosis is critical to prevent complications and minimise transmission. However, access to accurate diagnostic instruments is still problematic, especially in low-income regions.

Malaria diagnosis is usually performed by examination of thick and thin giemsa stained blood films where the laboratory technologists manually use microscopes to look for the malaria parasites (3, 4). Even though this approach is reasonably practical, it is rather time-consuming, somewhat subjective and dependent on qualified specialists. Some reasons include technician fatigue, poor imaging conditions, or variability in the blood smear preparation. Using artificial intelligence, specifically deep learning, to develop automated diagnostic tools can be an excellent solution to increase diagnostic efficiency and decrease the workload of healthcare professionals in limited resource settings (5).

Convolutional neural networks (CNNs), a type of deep learning, are very effective in the automated diagnosis of medical images. CNNs have been applied in many applications, including disease diagnosis, object identification, and segmentation (6). However, standard CNNs are data-hungry and labelled medical data are limited in many regions worldwide (7). To address these problems, this study uses the ConvNeXt architecture, which combines the ease of use of conventional CNNs with the hierarchical feature extraction of the latest models, such as vision transformers (ViTs).

The objectives of this study are as follows:

- To build a robust automated malaria diagnostic tool using the ConvNeXt architecture.
- To improve performance using transfer learning from pre-trained models on large datasets such as ImageNet.
- To investigate how data augmentation can make the model more robust in heterogeneous and low-resource situations.
- To evaluate the performance of ConvNeXt compared to other state-of-the-art architectures such as ResNet and Swin Transformer in terms of accuracy and computational efficiency.

In Figure 1, it is observed that the proposed ConvNeXt, which is a modernised convolutional neural networks architecture, outperforms other current complex models such as ResNet, DeiT, and Swin Transformer in both accurate training and computationally efficient when trained on ImageNet datasets (8). ConvNeXt applies to many fields due to its scalability and enhanced performance obtained through training on extensive datasets like ImageNet-22 (9).

**Figure 1:**
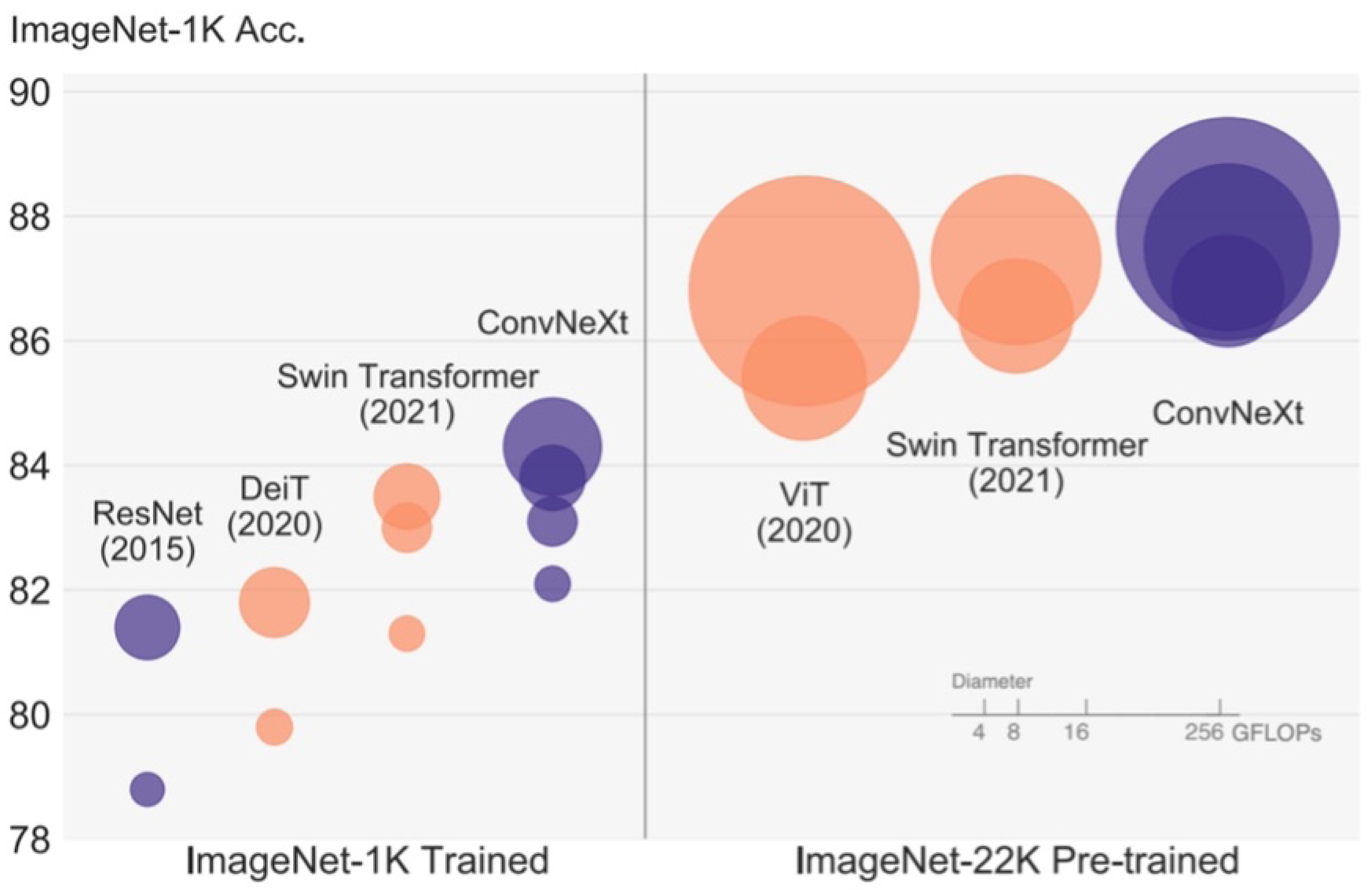
Performance of ConvNext on ImageNet. Source: (8)

Given the scarcity of big medical-labelled datasets, transfer learning and data augmentation have been extensively used in this work. Transfer learning is a technique that enables the models to use the knowledge learned from one task and apply it to another task, such as malaria detection from images. Conversely, the data augmentation technique enhances the dataset’s size by manipulating the image conditions to make the model less sensitive to practical variations.

This study explores a novel application of the ConvNeXt architecture, combining transfer learning with advanced data augmentation techniques for medical imaging. In contrast to conventional deep learning, feature extraction capabilities can be improved by transferring knowledge from the large-scale ImageNet dataset despite the relatively small dataset. Moreover, integrating explainable AI tools like LIME and LLaMA brings a unique dimension to a diagnostic process where the model’s decision-making can be visually and textually interpreted. These innovations highlight the opportunities for AI-driven systems to enhance diagnosis accuracy while promoting greater clinician acceptance by being transparent in AI predictions.

## 2. Materials and Methods

### 2.1 Dataset Acquisition

The dataset used for training and evaluating the deep learning models in this study was obtained from the Lister Hill National Center for Biomedical Communications (LHNCBC), part of the National Library of Medicine (NLM), which hosts a publicly available collection of malaria-infected blood smear images, available at (10). This dataset was initially collected at the Chittagong Medical College Hospital in Bangladesh, and the data are made up of thin blood smear images (11). The dataset comprises 27,558 images evenly distributed between parasitised and uninfected samples, with each category containing 13,779 images.

This balanced distribution ensures the reliability and validity of subsequent analyses and research findings (12). It prevents the model from being trained to overemphasise one class over the other in cases where the dataset could be more balanced between the two categories.

The blood smear images were photographed using a smartphone camera, which was held up to the eyepiece of a microscope; this configuration mimics the conditions likely to be found in low-resource settings. All the images were obtained from the blood smears stained with giemsa, the standard method of malaria diagnosis by microscopy. The images were then reviewed and labelled by expert technicians to determine whether or not the parasites were present. Each image had a dimension of 5312×2988 pixels, with the circular area depicted as the view through the microscope lens. As a result of the limited resources used in the imaging of blood smears and the variability in the preparation of the samples, the images had variations in lighting, contrast, and colour balance.

To avoid any possibility of identifying patients who may be reflected in the dataset, the patient data were de-identified before online publication. The Institutional Review Board (IRB) approved using the data at the NLM (IRB#12972)(11). Additionally, this study received ethical clearance from the University of Johannesburg’s School of Consumer Intelligence and Information Systems Research Ethics Committee (SCiiSREC) under ethical clearance code 2024SCiiS040. This clearance is valid for three years, starting on 1 August 2024. This approval ensures that the use of the data complies with the set ethical procedures for handling and analysing medical data.

In Table 1, the image counts are supplemented with both categories’ mean and standard deviation.

**Table 1:** Image Statistics for Parasitised and Uninfected Categories.

| Category | Image Count | Mean (R, G, B) | Standard Deviation (R, G, B) |
| --- | --- | --- | --- |
| Parasitised | 13,779 | [0.4507, 0.3882, 0.3955] | [0.3109, 0.2954, 0.2656] |
| Uninfected | 13,779 | [0.4478, 0.4841, 0.4634] | [0.2966, 0.3222, 0.2919] |

The mean and standard deviation values are pixel intensities, which are the extent of brightness or colour of a pixel on an image (13). The pixel intensity values are from red, green, and blue (RGB). These channels amount to the image’s colour, which ranges from 0, representing black, to 1, representing white. These are colour measurements for the pictures and consist of average colour intensities of the parasitised and uninfected samples regarding brightness and colour changes (14). As the value in a channel increases, the corresponding pixel intensity value will increase, implying a light and or intense colouration.

Pixel intensity analysis has been applied to detect parasites within microscopic images, particularly in identifying infected and uninfected cells. Studies such as (14) have shown the use of pixel intensities for classifying stained microscopy images based on colour variations. Similarly, several studies have demonstrated that pixel intensities are useful in diagnosing parasitic conditions (15–18). These works provide solid grounds for using pixel intensities, particularly in automated diagnostic systems.

The average pixel intensity of the colour channels in the “Parasitised” images is Red = 0.4507, Green = 0.3882, Blue = 0.3955. This indicates that the ‘Parasitised’ images have moderate pixel intensity levels in these channels. The standard deviations for the same set of images are approximately 0.3109, 0.2954 and 0.2656, indicating the pixels’ spread of intensity values. A larger standard deviation indicates greater dispersion; in this case, the dispersion is relatively moderate across the colour channels, which means there is some difference in image quality or staining intensity but not too extensive.

The pixel intensities for the “Uninfected” images are 0.4478, 0.4841, and 0.4634 in the three colour channels. These values show slightly higher pixel intensity, especially in the Green and Blue channels, than the “Parasitised” images. The standard deviations for the “Uninfected” images, which are 0.2966, 0.3222, and 0.2919, also indicate that the pixel intensity is moderately dispersed, similar to the “Parasitised” images but with variability across the channels. This dataset is well-balanced and statistically stable, which allows for the practical training of malaria detection models.

### 2.2 Image Pre-processing

#### 2.2.1 Image Resizing

The data used in this study were pre-processed before being applied to the deep-learning models used in this study. The input size of the original images at a resolution of 5312×2988 pixels offered an abundance of visual information; however, they were computationally costly and were not appropriate for the input dimensions of most current deep learning architectures. To this end, all images were resized to 224×224 pixels as most of the current ConvNeXt, Swin Transformer, and ResNet models have a standard input image size of 224×224 pixels (19).

These steps are essential, especially in resizing the images, because this has an impact on the speed with which the models can process the data; it lowers the memory usage during the process and allows the training of the models to be more efficient without losing much detailed information that is important in identifying malaria parasites. However, in most cases, when images are resized, they tend to lose some details, which can be crucial, especially when the resizing is performed to a great extent, such as here (20).

To avoid loss of information during image resizing, the Lanczos interpolation filter was used during the resizing of the image in Python, a method used for image scaling when the quality of the image should be preserved when downscaled (21, 22).

Lanczos interpolation is employed to resize the images while preserving the edges and fine details of the images by using the ‘sinc’ function so that the morphology of malaria parasites is well retained in the resized images (21, 23). Thus, applying this method, the details of the pre-processed images were preserved, which is crucial for accurately detecting and classifying the parasitised and uninfected blood smears by the deep learning models. Hence, despite the lower image resolution, which helped to enhance computational speed, the Lanczos filter was used to maintain an adequate image quality for the intended application.

Sinc or sinus cardinalis is a mathematical function applied in signal processing, especially in image processing for interpolation, especially in resampling or resizing images (24). The sinc function is defined as:

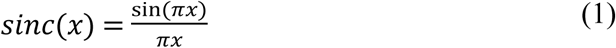

The sinc function, shown in Equation 1, is a continuous and periodic function defined by oscillations and decreases when moving away from zero (25). The sinc function is essential in Fourier analysis and can be described as the ideal low-pass filter in the frequency domain. It avoids the problem of aliasing, where different signals appear identical when sampled and produce distorted resampled images or signals.

This study uses the sinc function in Lanczos interpolation, which calculates new pixel colours while an image is being scaled. The sinc function decays and can be used as a weighting function when resampling surrounding pixel values, which produces much smoother transitions between pixels (26). This allows high-frequency details, such as sharp edges, to be kept when the image size is decreased, which is essential when dealing with the images of blood smears used in malaria detection.

#### 2.2.2 Image normalisation

After resizing the images, the normalisation process was performed to ensure that pixel intensities were consistent in the dataset. Normalisation is an essential procedure in deep learning since it enhances the stability of the learning process by preventing variations in the properties of the input data (27, 28). For the normalisation, the mean and the standard deviation of the pixel values of the images in each of the three colour channels, namely the red, the green and the blue, were computed.

The numerical results of the calculated mean values are listed in Table 1, are 0.4507 for the red channel, 0.3882 for the green channel and 0.3955 for the blue channel; the standard deviation values are 0.3109, 0.2954 and 0.2656, respectively. These values were then applied to normalise each image to ensure their pixel values had a mean around zero and the variance was one. This pre-processing step is essential to aid the models in learning and performing well when faced with data they have not encountered before by normalising the brightness and contrast of the images.

### 2.3 Data augmentation

Data augmentation is significant because it improves the transferability of the deep-learning models for malaria parasite identification (29). The lack of large, high-quality datasets is a common challenge in low-resource settings. Thus, data augmentation is useful in artificially increasing the amount of available data and introducing variability typical in real-life conditions (30, 31). To definite the models and make them capable of identifying the malaria parasites in varied situations, many data augmentation operations were performed on the base images, thereby creating a large dataset.

In this study, the techniques used for data augmentation were used for imitating diverse conditions under which blood smear images may be taken. Horizontal flipping applied the transformation that reflected the images across the x-axis, and vertical flipping was the transformation that reflected the images along the y-axis (32). These steps were critical to enable the model to learn how to identify malaria parasites, irrespective of their orientation. In the case of blood smear microscopy, the placement of the smears is only sometimes consistent.

Rotation is another transformation, where images were rotated at an angle of 45 degrees (33, 34). While preparing and analysing blood smears, they may not always be appropriately placed; hence, the model should be able to deal with rotated images. Scaling was used to control the size of the images with a scaling factor of 0.5 to 1.5 (33). This enhancement was functional in modelling the different magnification levels because the observed visual characteristics may range in size based on the microscope’s settings. This is because by training on scaled images, the model is in a better position to detect parasites regardless of their size.

Gaussian noise was added to the images to simulate potential noise observable in real-world image acquisition. The noise levels varied from 0.01 to 0.05 times the maximum pixel intensity value, which is 255 (33, 35). This made the training images similar to those taken in conditions that are not favourable, such as low light or inadequate focus of the camera. Contrast adjustments were made, whereby the contrast values were changed by a factor of 0.8 to 1.2 (33). Using contrast-enhanced images, the model could learn and distinguish parasites even in different image contrasts, thus improving its performance in different diagnostic conditions.

Besides these fundamental transformations, some more specific affine transformations were used, such as shearing (33), which changes the image along the x- or the y-axis to mimic some shifting or distortion that may occur while preparing the slides. Several techniques of blurring the images were used to make the images appear out of focus. Gaussian blurring with a sigma range of 0.0 to 3.0 was applied to the images (36). These techniques imitated the conditions of blurred images due to improper focus on the microscope, and thus, the model learned to recognise parasites in somewhat blurred images.

Sharpening was applied to the images to make the details more apparent, with the alpha range from 0 to 1 and lightness range from 0.75 to 1.5 (37). Sharpening improves the details in the images, especially the edges of the parasites, which are very important in identification. Colour-based augmentations, such as changes to hue and saturation, were also incorporated (38). These adjustments were informed by differences in staining method when preparing blood smears and differences in lighting or imaging systems. Elastic transformation and dropout were also applied, introducing more variability. The elastic deformations, which were controlled by alpha=50 and sigma=5, caused minor shifting of the image, and this was beneficial as the model could identify parasites even if there was a slight deformation in the image (39, 40).

Cropping was performed randomly by removing parts of the image with 0.02 to 0.1 pixels (41). This transformation aided in the model being more generalised so that it could still predict the images even if some parts of the images were cut or blurred. Channel shuffling was applied with a probability of 0.35, where the colour channels of the images were randomly interchanged to cope with colour channel variations (42).

Applying these transformations increased the dataset to 606,276 samples, where 303,138 is the number of parasitised samples and 303,138 is the number of uninfected samples. The mean pixel values for the parasitised images were proposed to be [0.44839746, 0.38788548, 0.39583275] while those of the uninfected images were [0.4479274, 0.4817927, 0.46273437]. The standard deviations were 0.30283427, 0.28804937 and 0.26085448 for the parasitised category, while those of the uninfected category were 0.2897945, 0.313396 and 0.285556. These statistics demonstrate the effects of the augmentation process that made the dataset more extensive and diverse in terms of the images’ features.

Figure 2 visually represents the dataset after data augmentation, explicitly comparing the number of images in two categories: The two main groups used in this study were parasitised and uninfected. There are two categories in total, each of them including about 303,138 images, proving the balanced distribution of the dataset after the augmentation process. This is very important in training the machine learning models, particularly in classification problems, to avoid being influenced by one category due to data sampling.

**Figure 2:**
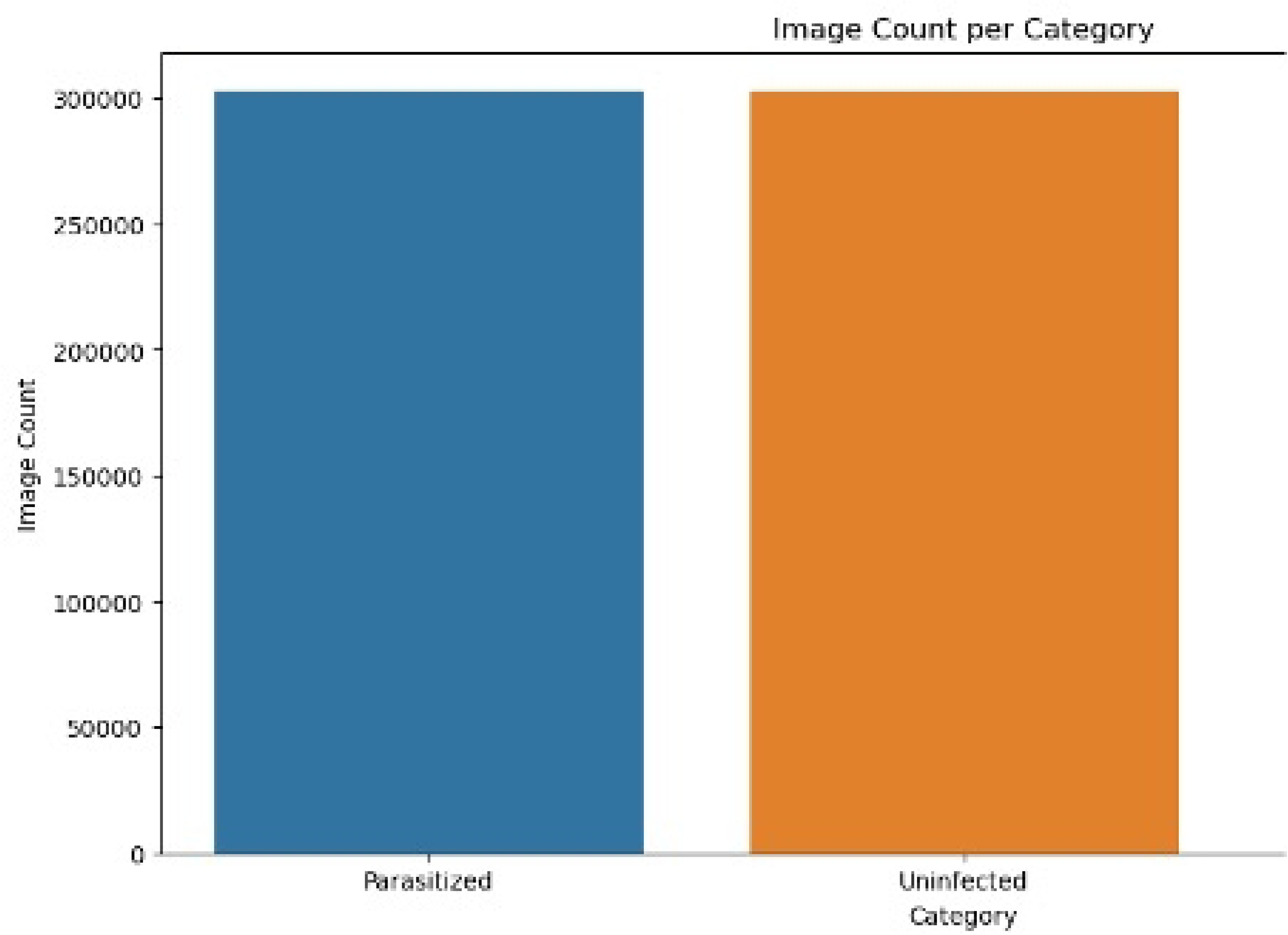
Image count per category (After data augmentation)

The augmentation made it possible to introduce variability in the form of geometric transformations, noise, scaling and colour variations, among others, as shown in Figure 3. This was performed by adding more training images and mimicking many scenarios one might encounter when capturing blood smears in practice, thus improving the generalisation capability of the models. Such augmentation aims to ensure that the models can identify the malaria parasites identifiable across various images and illumination and rotations, which are common in real-world applications.

**Figure 3:**
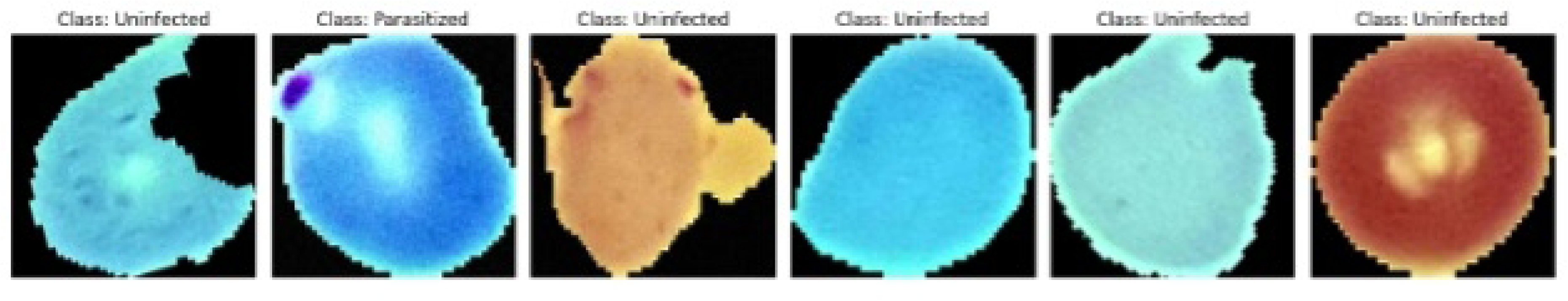
Sample images after data augmentation

Table 2 summarises the basic descriptive statistics of the characteristics of the augmented dataset used to train the malaria detection models. As seen in Table 2, the statistics of the dataset are reasonably even, and pixel intensity and variation are slightly different between Parasitised and Uninfected images, which will help the model during training. These colour distribution patterns are significant as they enable the model to distinguish between the parasitised and uninfected blood smear images, thus enhancing the model’s performance.

**Table 2:** Image Statistics for Parasitised and Uninfected Categories (After Augmentation)

| Category | Image Count | Mean (R, G, B) | Standard Deviation (R, G, B) |
| --- | --- | --- | --- |
| Parasitised | 303,138 | [0.44839746, 0.38788548, 0.39583275] | [0.30283427, 0.28804937, 0.26085448] |
| Uninfected | 303,138 | [0.4479274, 0.4817927, 0.46273437] | [0.2897945, 0.313396, 0.285556] |

### 2.4. Algorithms

This paper utilised several sophisticated deep-learning models to identify malaria parasites in microscopic blood smear images. These models cover various architectures that effectively capture local details, e.g., parasite shapes, and global information, e.g., cell distributions, in the images. The models used were the Swin Transformer (Swin Tiny), ResNet18, ResNet50, ConvNeXt Tiny, ConvNeXt V2 Tiny, and a modified version of ConvNeXt V2 called ConvNeXt V2 Remod. Every architecture has been chosen to work with high-resolution medical images and, at the same time, employ transfer learning, allowing the model to take knowledge from previously trained models trained on large datasets like ImageNet.

#### Swin Transformer Architecture

This study used the Swin Transformer, particularly the Swin Tiny model, to exploit its window-based multi-head attention (43). This architecture splits the input images into multiple non-overlapping local windows and then learns the self-attention from these local windows. This way, Swin Transformers can capture regional and global information in images, which is beneficial for tasks such as medical image analysis (44). The Swin Transformer architecture, as shown in Figure 4, is where images are divided into a sequence of non-overlapping patches. Then, a linear embedding layer is applied to generate patch tokens. This architecture is divided into four stages. Each stage consists of several Swin Transformer blocks and a patch merging layer, which reduces spatial dimensions while increasing the feature dimensions, allowing for both local and global context to be effectively captured across the entire image.

**Figure 4:**
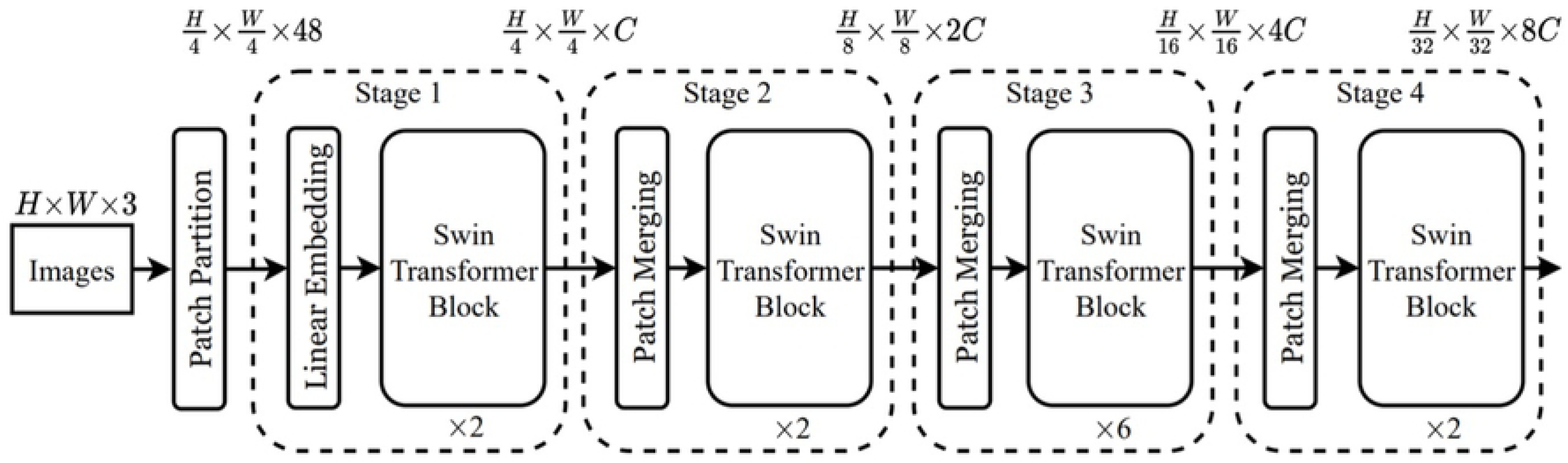
Architecture of a Swin Transformer. Source (43)

Mathematically, the self-attention mechanism within a Swin Transformer can be represented as:

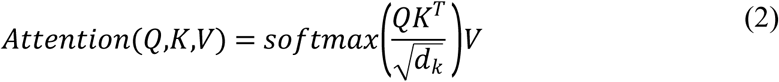

where Equation 2 represents the queries, keys, and values, respectively, and *d*_*k*_ is the dimension of the keys. The model incorporates a relative position bias term that enhances its ability to encode the spatial structure within each window.

Swin Tiny was pre-trained on ImageNet-1K, a dataset containing over a million labelled images. It achieved a top-1 accuracy of 81.2% and a top-5 accuracy of 95.5% on ImageNet, with 28 million parameters and a computation cost of 4.5 GFLOPs (45).

#### ResNet Architectures

The ResNet18 and ResNet50 were selected as the baseline models to be compared with the proposed models. These models are a part of the Residual Networks, which are the types of neural networks developed to address the vanishing gradient problem that is a big challenge in training deep neural networks (46). The residual block is central to ResNet’s architecture and is expressed mathematically as:

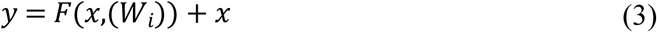

where x is the input, F(x,(W_i_)) is the learned transformation with weights W_i_ and y is the block output, as shown in Equation 3. This structure helps the network to learn identity mapping when deeper layers do not help enhance performance and allow a flow of gradients across many layers.

ResNet18 is a less complex model with 18 layers, while ResNet50 is a deeper model with 50 layers; hence, it can learn more complex data features. ResNet18 was pre-trained on ImageNet and fine-tuned for malaria detection, and the second model was pre-trained on ImageNet and fine-tuned for malaria detection (47). Although ResNet18 was used as the baseline model, ResNet50 had a deeper network and could thus identify more intricate visual patterns in the blood smear images and differences between them (48).

#### ConvNeXt and ConvNeXt V2 Architectures

ConvNeXt Tiny, a novel architecture based on conventional convolutional neural networks (CNNs), was another critical architecture used in this study (49, 50). Also based on Vision Transformers (ViTs), ConvNeXt is a model that combines the hierarchical architecture to incorporate both high and low-level features of images. Consequently, the feature extraction of Swin Transformer, ResNet, and ConvNeXt model block designs are different, as shown in Figure 5. Swin Transformer block captures local and global features with multi-head self attention (MSA) with shifted windows (w7×7), followed by layer normalisation and Gaussian Error Linear Unit (GELU) activation. A ResNet block is designed based on the residual structure of 1×1 and 3×3 convolutions, batch normalisation and Rectified Linear Unit (ReLU) activation to help feature learning. ResNet is modernised to ConvNeXt block by replacing 3×3 convolutions with depthwise convolutions (d7×7) with an extra parameter of channel multiplier, as well as using layer normalisation and GELU activation for better efficiency.

**Figure 5:**
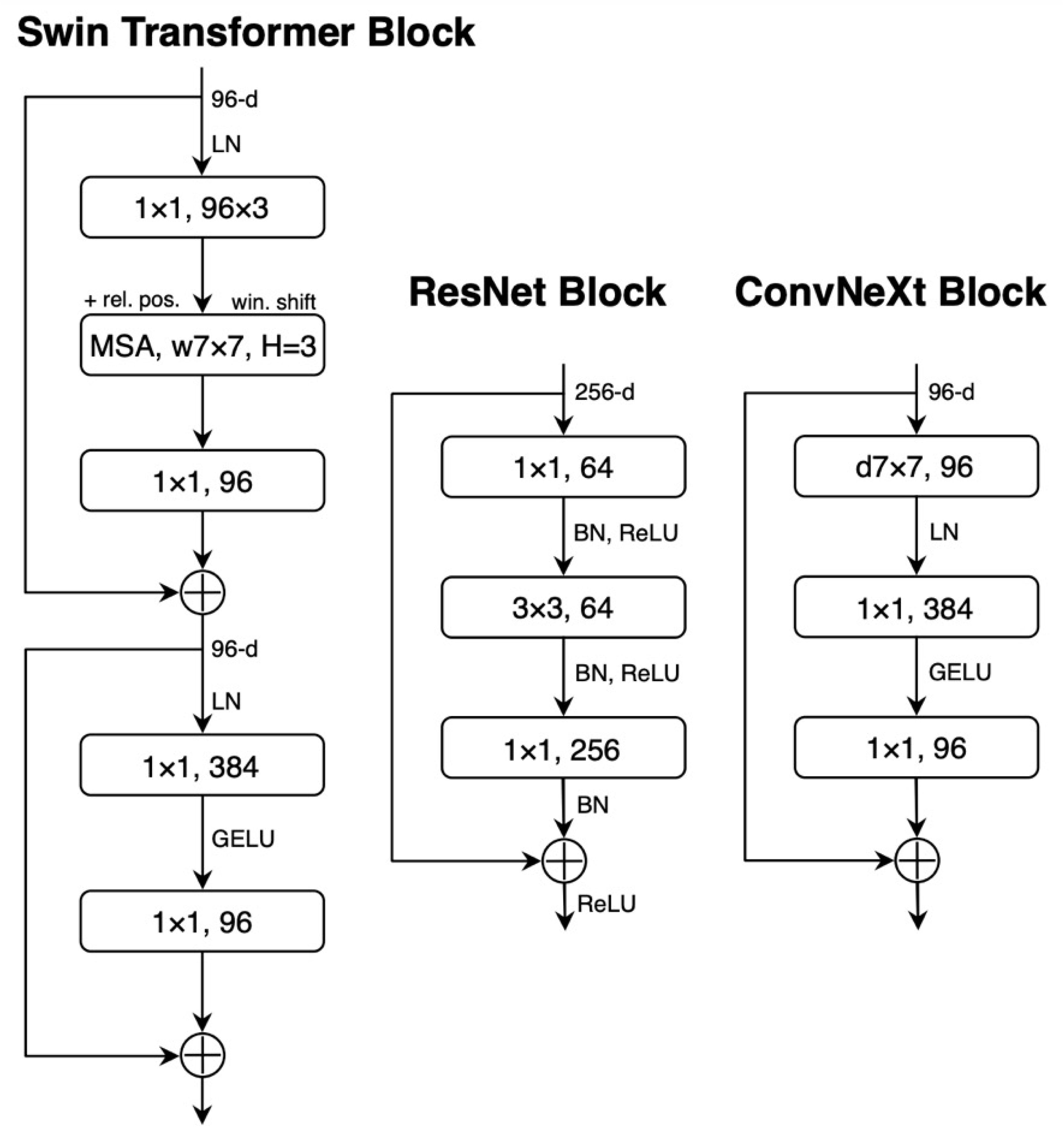
Block designs for the used models. Source (8).

The core operation in ConvNeXt is convolution, mathematically described as:

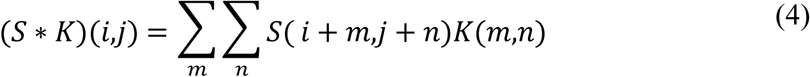

In Equation 4, S represents the input matrix (image), K, in Equation 4, is the convolutional kernel, and (i,j) denotes spatial positions in the image. ConvNeXt Tiny model was pre-trained on ImageNet, with the top-1 accuracy of 82.1% and has 28M parameters and 4.5 GFLOPs of computation. The pre-trained model was downloaded from GitHub and then fine-tuned to the malaria dataset to teach the model the features of detecting malaria parasites.

Following the success of ConvNeXt, ConvNeXt V2 had additional architectural enhancements, including learning rate scheduling and modified activation functions for improving the image classification task performance (50). The ConvNeXt V2 Tiny model employed in this study, which was also pre-trained on ImageNet, provided a top-1 accuracy of 83.0 per cent and had a computational complexity of 4.47 GFLOPs with 28. 6 million parameters. Similar to ConvNeXt V2, the model was fine-tuned on the malaria dataset to change the pre-trained weights of the model to adapt to the characteristics of the images of parasitised and uninfected blood smears.

#### ConvNeXt V2 Remod

This study’s ConvNeXt V2 Remod model was based on the ConvNeXt V2 Tiny architecture. In the training process, label smoothing was used with α=0.1 (51, 52). Label smoothing shifts the target labels to prevent the model from attaining extreme confidence and helps deter the overfitting of the model (52). Mathematically, label smoothing modifies the loss function as follows:

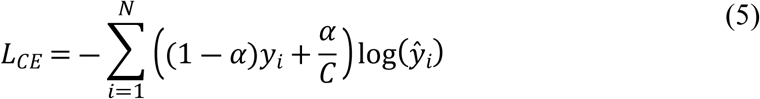

where C is the number of classes (in this case, C=2 for binary classification), alpha distributes a small part of the probability mass to the incorrect classes, as shown in Equation 5.

The block structures of ConvNeXt V1 and V2, shown in Figure 6, show the core improvements made from one version to the other. The ConvNeXt V1 block consists of depthwise convolutions (d7×7) and then layer normalisation (LN) and GELU activation for efficient feature extraction. The first version of the model introduced LayerScale, which helped stabilise the process. On the other hand, the ConvNeXt V2 block keeps the heart of V1 but introduces Generalised ReLU Normalisation (GRN). This new state-of-the-art normalisation technique enhances the model’s stability and efficacy. This addition of GRN with the elimination of LayerScale boosts the generalisation ability of ConvNeXt V2 across several tasks.

**Figure 6:**
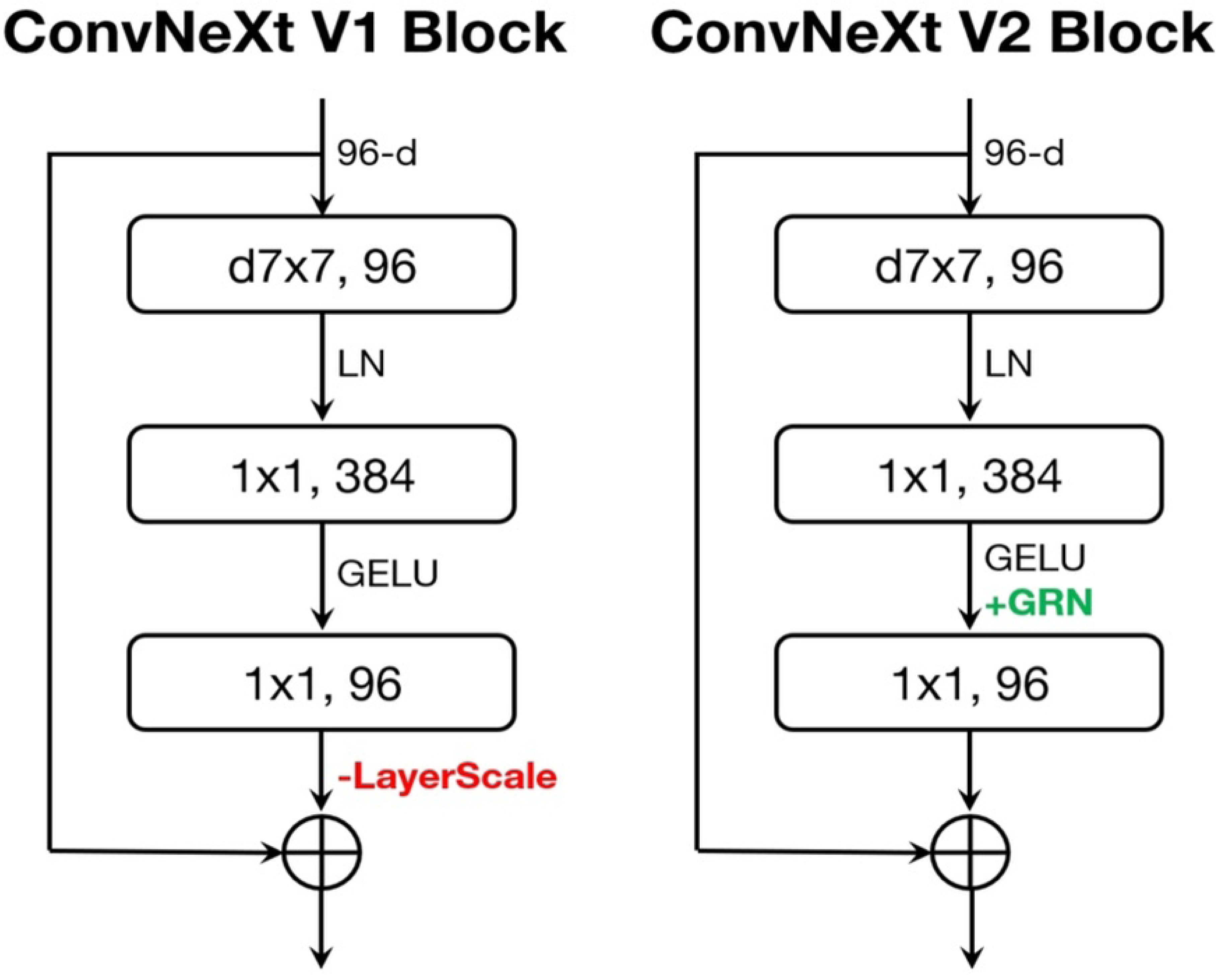
ConvNeXt block designs. Source (50).

The AdamW optimiser, shown in Equation 6, was selected for this task as it is a version of the Adam optimiser with weight decay for better generalisation and reduced overfitting (53, 54). AdamW updates the model’s weights using first- and second-order moments of the gradients.

The main distinction between Adam and AdamW is that in AdamW, the weight decay is not applied directly to the gradients, which allows the magnitude of the weights to be preserved.

The learning rate was set to 0.0005, and a weight decay of 0.01 was applied to penalise large weights, thus avoiding overfitting the training data (55). The AdamW optimiser is defined as:

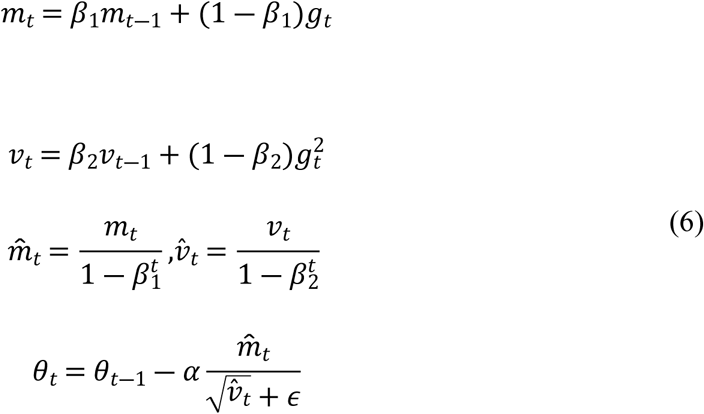

where:

g_t_ is the gradient of the loss with respect to the parameters at step t

m_t_ and v_t_ are the moving averages of the gradient and its square, respectively,

theta represents the model parameters at step t

alpha is the learning rate, and

Beta 1, Beta 2 and epsilon are hyperparameters of the optimiser.

To improve learning efficiency, the OneCycleLR scheduler was applied (56). This scheduler modifies the learning rate throughout training to balance exploration (high learning rate) and refinement (low learning rate). The learning rate is first set to maximum value and then decreases over time, which helps the model avoid local minimum in the initial training phase while fine-tuning the weights when training continues.

This is set to zero initially, rises to a maximum of 0.0005 and then decreases using a cosine annealing schedule, shown in Equation 8, over ten epochs. The cosine annealing formula is given as Equation 7 by:

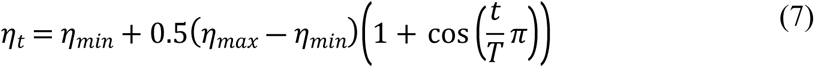

where:

Eta t is the learning rate at time step t,

Eta max and Eta min are the minimum and maximum learning rates, and

T is the schedule’s total number of time steps (epochs).

To optimise the training of the NVIDIA^®^ Tesla^®^ P100 graphics processing unit (GPU) used in Kaggle, PyTorch’s automatic mixed precision (AMP) was implemented to train in mixed precision. It is a technique where half of the computations are performed in a 16-bit floating-point format, whereas the other half are performed in a 32-bit floating-point format. The gradients are calculated in the FP16 format for the sake of optimisation, while the master weights are stored in the FP32 format for the sake of numerical accuracy.

For this, gradient scaling was used to deal with the differences between FP16 and FP32. Gradient scaling is the process of scaling the gradients by a specific factor to prevent the gradients from being too small (and thus causing underflow) or too large (which will cause overflow). This scaling assists in controlling the training process, especially for deep networks with numerous parameters to be estimated.

The training of the ConvNeXt V2 Remod model was performed for ten epochs, with the learning rate being adjusted in a way that aims at deriving the highest performance. The model was trained with high performance and reliability using mixed precision through automatic mixed precision and with the help of checkpointing. Thus, the optimisation strategies and the advanced architectural features helped to enhance the recognition of malaria parasites in blood smear images at the end of the training process.

### 2.5 Model Development

#### 2.5.1 Transfer learning

This work applied transfer learning, where pre-trained models were obtained from GitHub repositories. Transfer learning offered several advantages as it significantly cut down training time. As the models had previously been trained on large datasets such as ImageNet, the computational power and time needed for fine-tuning the models for malaria classification were considerably lower than that required for training new models. Secondly, transfer learning played a role in reducing overfitting, which is a big problem when dealing with limited samples. Through pre-training, the models with a large set of images, continued to perform well on the malaria classification task due to their generalisation ability.

In addition, transfer learning was used to ensure the models’ accuracy and robustness were not compromised by the new task of distinguishing between parasitised and uninfected blood smears since the models could still rely on their prior experience with identifying general objects in images (57). This approach was instrumental in resource-limited settings where access to vast amounts of labelled data and powerful computing resources is limited.

Swin Transformers, ResNet 18, ResNet 50, ConvNeXt Tiny and ConvNeXt V2 Tiny were the models employed in this study, all pre-trained on large datasets of generic images. There are several benefits of using these pre-trained models; first, they are already equipped with feature extraction capabilities that can be fine-tuned for a specific task, for instance, malaria parasite detection in microscopic blood smear images (58–60).

The pre-trained Swin Transformer models were acquired from the repository managed by Microsoft and are accessible at (61). Swin Transformers are well-suited for handling high-resolution images due to their unique mechanism of dividing images into non-overlapping windows, enabling efficient computation of self-attention within each window (43, 62). This approach makes it possible for the model to learn both the image’s local and global features, which is very important during the classification, especially in tasks such as parasite identification.

ConvNeXt and ConvNeXt V2 models used in this work were obtained from the public repositories established by Facebook AI Research. ConvNeXt models can be accessed at (63). They represent advancements over regular convolutional neural networks (CNNs) with features derived from Vision Transformer architectures (8, 49). ConvNeXt V2, accessible at (64), incorporates improvements that include better resource management and improved feature extraction and, hence, is even more suitable for tasks such as malaria classification (50).

These models were first pre-trained on ImageNet, one of the most commonly used datasets in the computer vision field (65). The ImageNet dataset is one of the most extensive, with over 14 million labelled images spanning 1,000 categories of objects, including animals, buildings, nature, and food, amongst others (66). This large amount of data helps these models to learn the features well. Hence, they are ideal for transfer learning applications, such as detecting malaria parasites with limited datasets.

Fine-tuning these models for classifying parasitised and uninfected blood smears involved adjusting the final fully connected layers to output two classifications: The blood smears were either parasitised or uninfected (67). It is essential to replace the pre-trained models’ last layers with new layers specifically designed for malaria parasite detection. While the pre-trained models are well-trained in the general characteristics of the images, such as edges, patterns, and textures, the final layers can be focused on learning the unique characteristics of the blood smears relevant to malaria diagnosis. In this case, the final layers were replaced with custom layers designed for binary classification, parasitised versus uninfected.

#### 2.5.2 Training Procedure

The training process for this study was performed on Kaggle, which offered the use of NVIDIA^®^ Tesla^®^ P100 GPU. This is mainly because of the GPU’s architecture and computational ability for deep learning. The Tesla^®^ P100 is based on NVIDIA^®^’s Pascal architecture, includes 16GB of HBM2 memory and performs 10.6 TeraFLOPS in single-precision floating-point operations per second. Such a computational capability helped handle large image datasets used in this study without experiencing low speeds. The P100 had enough memory to operate on large and high-resolution images so that batch processing of models could be performed faster during training.

The training was performed using mixed-precision training with the help of PyTorch’s automatic mixed precision (AMP) (68). AMP is a technique which enables deep learning models to train faster and with less memory than standard training (68, 69). AMP works on some parts of the network. For instance, the convolutions are computed using 16-bit floats, while the rest of the model’s weights and gradients are calculated using 32-bit floats to balance between speed and accuracy.

This technique also enhances the speed of training and optimisation of the GPU, especially the Tesla^®^ P100, which performs operations on large datasets with great accuracy. Furthermore, the vanishing and exploding gradient issues that are evident when training deep networks with vast parameter spaces like ConvNeXt or Swin Transformer models are also solved by AMP.

##### Optimisation Strategy

This study opted to use the Adam optimiser, as shown in Table 3, which has been known to be efficient in adjusting the learning rate during training (70). Adam uses the benefits of AdaGrad and RMSProp optimisations involving gradient averages and second-order moments to adjust the model weights (53, 71). This is advantageous for Adam for handling the sparse gradients and noisy data, which are familiar with medical image data. The learning rate of the optimiser was set to 0.0001 for most models, including ConvNeXt Tiny, ResNet18, and ResNet50, due to its ability to fine-tune deep learning features, especially for medical imaging data that typically contain noise and sparse gradients (72). A low learning rate is essential to guarantee that the model gives only small weight adjustments to make the best estimate without overfitting highly sensitive models on the loss surface (73). For the ConvNeXt V2 Remod model, a higher learning rate of 0.0005 was used alongside weight decay of 0.01 to improve model generalisation (74).

**Table 3:** Hyperparameters used for fine-tuning different models.

| Model | Learning Rate | Batch Size | Optimiser | Weight Decay | Loss Function | Scheduler | Epochs | Mixed Precision |
| --- | --- | --- | --- | --- | --- | --- | --- | --- |
| ConvNeXt Tiny | 0.0001 | 128 | AdamW | Not Used | Cross Entropy Loss | None | 10 | GradScaler & autocast |
| Swin Transformer | 0.0001 | 128 | Adam | Not Used | Cross Entropy Loss | StepLR | 10 | GradScaler & autocast |
| ResNet18 | 0.0001 | 32 | Adam | Not Used | Cross Entropy Loss | None | 10 | GradScaler & autocast |
| ResNet50 | 0.0001 | 128 | Adam | Not Used | Cross Entropy Loss | None | 10 | GradScaler & autocast |
| ConvNeXt V2 Tiny | 0.0001 | 128 | AdamW | Not Used | Cross Entropy Loss | None | 10 | GradScaler & autocast |
| ConvNeXt V2 Remod | 0.0005 | 128 | AdamW | 0.01 | Cross Entropy Loss (Label Smoothing = 0.1) | OneCycleLR (Cosine) | 10 | GradScaler & autocast |

The adaptive learning rate of Adam makes the model converge faster than the normal stochastic gradient descent (SGD) (75). In this study, it was beneficial for large and elaborate models such as ConvNeXt and Swin Transformer with many parameters. Adam allowed the model to automatically and adaptively set the learning rate for each parameter, which made the model optimise itself and increase the training process’s convergence rate.

##### Gradient Scaling and Stability

Gradient scaling was used during the optimisation to avoid training process instabilities (76). It is a method applied in mixed-precision training to prevent the gradients from becoming too small (vanishing gradients) or too large (exploding gradients) (77). In large networks, such as those used in this study, the parameter space is also significant, which can result in oscillations during training. Applying gradient scaling before backpropagation helped avoid the potential numerical precision problems common in deep learning models.

A learning rate scheduler was used to stabilise the training process during implementation. A learning rate scheduler adjusts the learning rate during training according to the given performance of a model (78, 79). In this case, the scheduler had a learning rate reduction of 0.1 every seven epochs if the validation loss did not decrease. This technique is very helpful in preventing the model from becoming trapped in the local minima. When the validation loss stalls, reducing the learning rate helps the optimiser fine-tune the model weights and allows the model to perform better and converge. For ConvNeXt V2 Remod, a OneCycleLR (cosine) scheduler was applied to balance fast convergence with the risk of overfitting

##### Loss Function: Cross-Entropy Loss

The loss function used for this study was the Cross-Entropy Loss function, which is most suitable for the binary classification of the malaria parasite cases (parasitised and uninfected red blood cells) (80). Cross-entropy loss measures how far the predicted class probabilities are from the actual class labels (81). It is more useful in classification as it penalises the predicted output that is not in line with the actual output. In the case of ConvNeXt V2 Remod, label smoothing was also applied to the cross-entropy loss to mitigate the effect of noisy labels and improve generalisation. In this work, the model was trained to predict the likelihood of a given blood smear image being parasitaemic. The output from the final layer in the model is a vector of predicted probabilities, which are compared to the actual class labels, and the weights of the model are updated to minimise this loss function known as Cross-Entropy Loss.

Mathematically, Cross-Entropy Loss is computed as:

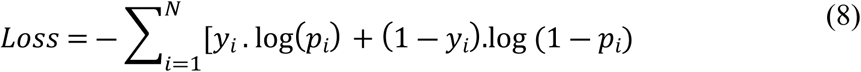

where:

N is the number of samples,

y_i_ is the true label (either 0 for uninfected or 1 for parasitised),

p_i_ is the predicted probability for the corresponding class.

In this context, Cross-Entropy Loss, displayed in Equation 8, helps the model to give a high probability to the correct class and a low probability to the incorrect class (82). Since the dataset has equal parasitised and uninfected images, Cross-Entropy Loss prevents the model from leaning towards one class and produces equally good results.

### 2.6 Model Application and Deployment

To enhance the applicability of the trained ConvNeXt models, a web application was developed using Gradio to facilitate real-time identification of malaria parasites in blood smear images. The Python library Gradio designed application has an interface where users can upload blood smear images and receive real-time diagnoses in a limited resource environment. It is accessible at (83).

The application (app) was designed with two main functionalities: The proposed solution includes image classification using ensemble models and the explanation of the predictions using the LIME model (Local Interpretable Model-agnostic Explanations) and Llama 3.1 by Facebook Research (84–87).

The application presents novel contributions through the use of an ensemble of deep learning models (ConvNeXt Tiny and ConvNeXt V2) that help improve diagnosis performance and reliability. To do this, the app takes the average of these models’ predictions by assigning weights to each of the models to make a more accurate determination on whether a blood smear is parasited or not. This ensemble approach outperforms the conventional single-model systems employed in medical diagnostics to provide a more accurate result, especially in the limited resource environment (88). Furthermore, the app employs mixed-precision training with the help of GradScaler, which ensures the high performance with the minimal consumption of resources, which will be beneficial for the areas with poor infrastructure.

One of the main aspects of the app is the explainability element, using LIME for visualisation and LLaMA for textual elaboration (87, 89). LIME gives out visual maps that demonstrate the parts of the image which are used in making the decision, which helps in improving the explainability of the predictions to medical practitioners (89). LLaMA enables an additional step in explaining the results as it generates human-readable descriptions of the context and, therefore, helps interpret the machine’s predictions (87). These explainability features together with efficiency in the use of resources make the Malaria Diagnosis App not only unique but also very useful for use in malaria endemic areas.

#### 2.6.1 Use of best performing models

The application combines the strengths of two fine-tuned models, namely ConvNeXt Tiny and ConvNeXt V2 Tiny Remod, through the ensemble method (90). This approach combines the prediction of the first model with the second model’s prediction to make the final decision, increasing the overall reliability of the decision made (91). From the two models used in this study, ConvNeXt Tiny was purposely selected for its computation efficiency, which allows it to process images at high speed. At the same time, ConvNeXt V2 Tiny Remod was chosen because of its higher accuracy and precision, as highlighted in the comparative analysis.

Every image the user uploads is first processed through a pre-processing step, which includes resizing the image to 224 x 224 pixels and normalising the image using the mean and standard deviation of the malaria dataset. This check helps meet the conditions the trained models expect as input data. The image is then passed through both models, and the average output of the two models is computed. The final decision on whether a blood smear is parasitised or uninfected is based on the average of the two models presented in this work. This approach enhances the diagnostic performance and decreases the rates of false-positive results, thus making the diagnosis more accurate.

#### 2.6.2 Model Explanation with LIME

To further enhance the transparency and interpretability of the malaria parasite detection models, the LIME (Local Interpretable Model-agnostic Explanations) algorithm was employed. LIME works by perturbing the input data - in this case, the blood smear images - and observing how the model’s predictions change in response to these perturbations. The algorithm generates explanations by locally approximating the model’s decision boundary and identifying the regions within the image that most influence the final classification. This process is precious for understanding deep learning models, often called “black boxes.”

Mathematically, LIME can be understood as follows:

Using a kernel function, LIME approximates the complex model around the local region of the explained instance: *π*(*x*_0_, *x*^′^).

LIME minimises the following loss function to generate explanations:

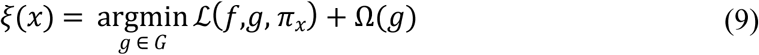

where:

Xi(x) is the explanation for the instance x,

G is the class of interpretable models, L(f,g,Pi_x_), in Equation 9, is the loss function that measures the fidelity of the interpretable model g to the complex model f in the local region defined by the kernel Pi_x_.

Omega(g) is a complexity term that penalises the complexity of the interpretable model g, ensuring that the explanation remains simple and human-readable.

The kernel function Pi(x_0_,x’) assigns higher weights to perturbed samples closer to the original instance x_0_ ensuring that the explanation focuses on the local behaviour of the model. In the case of image classification, this kernel function is often defined based on the Euclidean distance between the perturbed image *x*^′^and the original image x_0_.

Once the simpler model g(x) is trained to highlight the regions in the image most responsible for the model’s prediction. These highlighted regions correspond to the image features—such as ring-shaped structures or abnormal cell morphologies that the model associates with malaria parasites.

For each blood smear image analysed, LIME produces a visual heatmap highlighting the most influential regions in the classification decision. These highlighted areas allow healthcare professionals to understand why the model classified an image as parasitised or uninfected. This feature is crucial in clinical settings because it provides clinicians with tangible visual cues to validate the model’s predictions. It bridges the gap between complex machine learning models and human interpretability, making AI-driven diagnostic systems more transparent and trustworthy.

By offering healthcare professionals clear, visual evidence of what the model “sees,” LIME enhances decision-making support. Physicians and laboratory technicians are given the output of the model and the rationale behind the decision, building trust in the diagnostic process. Moreover, the mathematical framework behind LIME ensures that the explanations are robust and focused on the most significant aspects of the model’s behaviour.

#### 2.6.3 Integration with LLaMA for Diagnostic Insights

The above outputs were complemented with textual descriptions produced by the Large Language Model Meta AI (LLaMA) language model to provide more specific case information (87). It enabled the app to classify blood smear images and briefly describe each classification based on context. An advantage of the system was that it used a pre-trained LLaMA model to provide interpretations in simple language of why a particular classification of the image as parasitised or uninfected was made.

In the case of blood smears classified as parasitised, the LLaMA model offered some understanding of the visual cues that contributed to this classification. It also showed the presence of trophozoites, the asexual form of malaria parasite presenting as ring-shaped structures within red blood cells and some other abnormalities, including irregular shapes of the red blood cells. When these features were identified by the ConvNeXt models, they marked a critical point in signalling an infection, which LLaMA translated into a layman’s understanding.

The use of natural language processing in combination with machine learning algorithms has its advantages in clinical settings as it helps make the results more understandable. This way, the system expands the original AI’s predictions with natural language descriptions to fulfil the requirements of healthcare workers. Extending the ability to justify the prediction made by the model helps enhance the credibility of the model’s predictions. It enables the clinician to act on the insights provided by the AI system more effectively. Ultimately, this integration makes the system more practically beneficial for diagnostic pipelines.

#### 2.6.4 Deployment

To develop the application, a Gradio interface with a well-organised and easily navigable layout was employed to facilitate the uploading of microscopic blood smear images with the subsequent immediate diagnostic output. When an image is uploaded, the system processes the image through a pre-processing, classification, and explanation phase. The results are presented to the user in both textual and graphical formats. It also builds on the features of Gradio’s interface that enhance the interaction, especially for individuals who may not be quite conversant with the software. After the image has been uploaded, the fine-tuned ConvNeXt models are applied to the image, and the output is either parasitised or uninfected. In addition, a LIME model visualises the image regions that led to the classification decision.

The additional textual explanation that LLaMA can help practitioners understand the outcomes more straightforwardly. This dual output, which is both visual and textual, significantly improves the understandability of the AI-based differential diagnosis. For this reason, the app has a supporting backend script that controls memory usage, especially in environments with constrained resources. It is the same whether the code runs on a GPU for fast computations or a CPU in less powerful environments; the app backend is designed to be non-memory bound.

This design helps the app work effectively even in regions with limited resources, which is typical for malaria-endemic areas. Since the app was developed based on Gradio, it has a high level of adaptability. It can be adapted for mobile versions or integrated into existing healthcare systems. This portability is especially valuable in malaria-endemic areas where portable diagnostic tools are crucial for front-line healthcare providers. With this app accessible online, installed on mobile devices or in healthcare systems of a specific region, the lack of time and AI in detecting malaria becomes relative, thus increasing the chances of curing patients in areas where malaria still needs to be solved.

## 3. Results

In this study, deep learning has been widely used for malaria parasite detection, and many evaluation metrics have been used to assess the performance of the models in each of the classification tasks. To evaluate the performance of the selected models such as Swin Tiny, ResNet18, ResNet50, ConvNeXt Tiny, ConvNeXt V2 Tiny and a re-modified version of ConvNeXt V2 Tiny, various criteria were used. These metrics are accuracy, precision, recall, F1 score and ROC-AUC (Receiver Operating Characteristic – Area Under the Curve). Relative measures, including log loss, MCC, specificity, balanced accuracy, Cohen’s kappa, G-mean, FPR and FNR, were also used to assess the discriminative ability of the models between parasitised and uninfected blood smears.

The basic measure is accuracy, in Equation 10, and is calculated as the number of correctly classified observations divided by the total observations. Mathematically, accuracy can be expressed as:

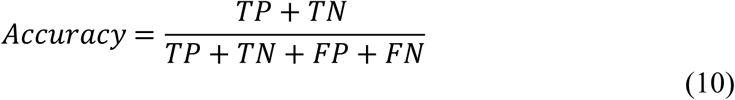

The symbols used are TP for True Positives, TN for True Negatives, FP for False Positives and FN for False Negatives. Since the dataset used in this study was balanced, accuracy is a good measure to use, although it may not be the best measure for imbalanced data. Both precision and recall, thus, give a more detailed picture.

Precision or Positive Predictive Value, shown in Equation 11, is the probability that a positive identification is correct. It is calculated as:

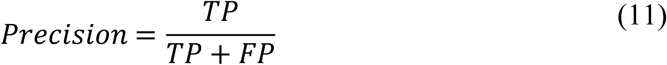

Precision is crucial when false positives are to be avoided, for example, in a diagnosis where a wrong result can mean a patient is subjected to a treatment he does not need.

Recall, or Sensitivity, is the ability to find all the positives out of the cases that are in fact positive. It is calculated as:

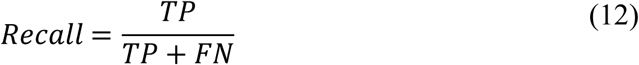

A high recall rate, calculated as shown in Equation 12, means that the model accurately predicts most of the positives, which is especially important for tasks that involve risk, such as diagnosing malaria, when failure to identify a positive case (false negative) may have severe outcomes.

The F1 score, in Equation 13, integrates precision and recall into a single score that considers both of them equally profitable or unprofitable. The F1 score is defined as the harmonic mean of precision and recall:

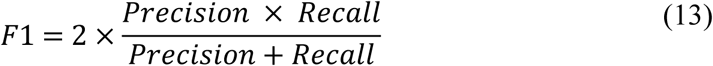

This metric gives us a more holistic picture of the model’s performance when it needs to balance trade-offs between precision and recall.

The ROC-AUC was also considered in this study, as shown in Equation 14. The true positive rate (TPR) is plotted against the false positive rate (FPR), giving the ROC curve, which graphically represents how the model is discriminatory. This performance is summarised by the AUC (Area Under the Curve) where the AUC score of 1 represents perfect discrimination and of 0.5 represents no discrimination. The AUC can be mathematically expressed as:

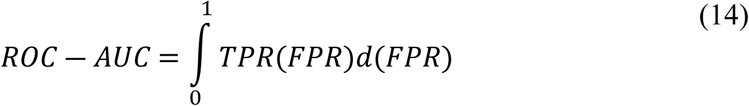

A particularly useful metric for model performance in medical diagnostics is ROC AUC, whose purpose is to evaluate model performance across different thresholds and, thereby, across a range of decision boundaries.

Furthermore, uncertainty of predictions was measured using log loss, in Equation 15. This metric is a rigorous metric because it penalises confident incorrect predictions more heavily than less confident ones. Mathematically, it is defined as:

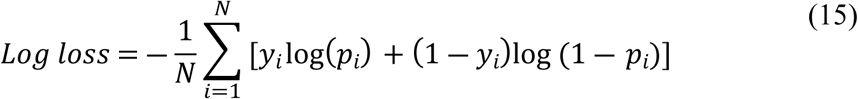

where *y*_*i*_ is the actual label and *p*_*i*_ is the predicted probability for the positive class.

Quality of binary classifications was measured using Matthews correlation coefficient (referred to as MCC), given as Equation 16. It covers all four confusion matrix categories (TP, TN, FP, FN) and works well in imbalanced dataset cases, even though, the dataset used in this particular case was balanced. MCC is calculated as:

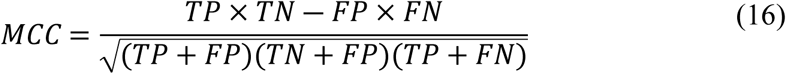

True Negative Rate, OR Specificity, is the number of actual negatives identified as negatives. Recall addresses positive cases, and it is complemented by it. Agreement between the model’s predictions and true labels were also measured using Cohen’s kappa, refined by chance, and G-mean that is a geometric mean of sensitivity and specificity.

Moreover, using both False Positive Rate (FPR), in Equation 17, and False Negative Rate (FNR), in Equation 18, gives additional information about what type of errors the models made. FPR, calculated as:

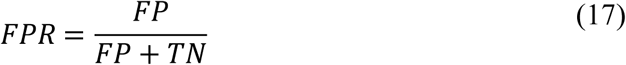

indicates how often the model incorrectly labels uninfected images as parasitised, whereas FNR, calculated as:

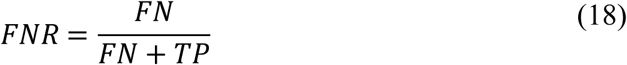

reflects how often the model fails to detect actual parasitic infections.

Every metric has its advantages and disadvantages. For instance, while accuracy is easy to understand and calculate, it can be misleading with imbalanced datasets, though that was not an issue in the dataset used in this study. Precision is relevant in situations with the least tolerance for error in predictions. At the same time, recall is important in situations requiring detecting as many positive cases as possible. The advantage of the F1 score is that it is a balanced metric, but it can be less useful when precision and recall are much different. ROC-AUC is a robust measure in different thresholds. However, it can be sensitive to the presence of imbalanced data; this was not a problem in this work. The log loss ensures a model is not overconfident in its predictions, and MCC provides a more complementary score for performance, especially when dealing with imbalanced datasets. FNR and G-mean increase the understanding of the model’s failure modes. Tables 4.1, 4.2 and 4.3 present model performance comparisons across these critical metrics.

**Table 4.1.** : Model performance comparison across critical metrics.

| Model | Accuracy | Precision | Recall | F1 Score | ROC-AUC |
| --- | --- | --- | --- | --- | --- |
| Swin Tiny | 0.613988 | 0.572842 | 0.896420 | 0.699000 | 0.679830 |
| ResNet18 | 0.625687 | 0.572930 | 0.987379 | 0.725112 | 0.839727 |
| ResNet50 | 0.813671 | 0.731378 | 0.991502 | 0.841803 | 0.951111 |
| ConvNeXt<br>Tiny | 0.958646 | 0.944025 | 0.975110 | 0.959316 | 0.991124 |
| ConvNeXt V2<br>Tiny | 0.544330 | 0.523228 | 0.998578 | 0.686663 | 0.815418 |
| ConvNeXt V2<br>Tiny Remod | 0.981249 | 0.979453 | 0.983123 | 0.981285 | 0.996633 |

**Table 4.2:** Model performance comparison across critical metrics.

| Model | Log Loss | MCC | Specificity | Balanced<br>Accuracy |
| --- | --- | --- | --- | --- |
| Swin Tiny | 1.402099 | 0.276272 | 0.331555 | 0.613988 |
| ResNet18 | 1.681094 | 0.364074 | 0.263995 | 0.625687 |
| ResNet50 | 0.831209 | 0.671230 | 0.635839 | 0.813671 |
| ConvNeXt Tiny | 0.116142 | 0.917790 | 0.942181 | 0.958646 |
| ConvNeXt V2<br>Tiny | 2.224143 | 0.212159 | 0.090081 | 0.544330 |
| ConvNeXt V2<br>Tiny Remod | 0.099783 | 0.962506 | 0.979376 | 0.981249 |

**Table 4.3:** Model performance comparison across critical metrics.

| **Model** | **Cohen's**<br>**Kappa** | **G-Mean** | **FPR** | **FNR** |
| --- | --- | --- | --- | --- |
| Swin Tiny | 0.227975 | 0.545172 | 0.668445 | 0.103580 |
| ResNet18 | 0.251374 | 0.510552 | 0.736005 | 0.012621 |
| ResNet50 | 0.627341 | 0.794000 | 0.364161 | 0.008498 |
| ConvNeXt Tiny | 0.917292 | 0.958505 | 0.057819 | 0.024890 |
| ConvNeXt V2<br>Tiny | 0.088659 | 0.299922 | 0.909919 | 0.001422 |
| ConvNeXt V2<br>Tiny Remod | 0.962499 | 0.981248 | 0.020624 | 0.016877 |

The ConvNeXt models, especially the ConvNeXt V2 Tiny Remod, were highly able to differentiate between parasitised and uninfected samples with an accuracy of 98.1%, as shown in Table 4.1. This means the model can differentiate between malaria and other samples with high accuracy and low chances of misclassification, making it suitable for real-world applications. However, in the case of the Swin Tiny model, the accuracy was relatively low, standing at 61.4%, significantly indicating some discrepancies in the classification when differentiating between the two types of samples, which may raise a question regarding its practicability for real-time diagnosis.

Another critical factor in measuring the models’ performance is their capability to minimise the number of false positives. Here, the accuracy of the ConvNeXt V2 Tiny Remod was impressive, with a precision of 97.9%, as shown in Table 4.1. This high precision shows that the model made only a few mistakes when identifying parasitised samples, thus reducing the possibility of unnecessary treatment for unparasitised ones. On the other hand, the precision of Swin Tiny stands at 57.3%, which is an indication of the model’s propensity to classify healthy samples as infected ones. Such high false positive readings can result in mistreating patients and unnecessary procedures in a clinical context.

The ability of the model to correctly predict actual malaria cases is also important, as measured by recall. ConvNeXt V2 Tiny Remod had a strong recall, achieving 98.3% in this task. A recall of 98.3% means that the model failed to identify only 1.7% of the parasitised cases, which shows that the model has high sensitivity in detecting malaria-infected blood smear image. Nevertheless, a higher recall of 89.6% was observed from Swin Tiny, which shows that the model correctly identified a large number of parasitised samples; however, it also had a low precision, meaning that Swin Tiny often misclassified uninfected samples as parasitised.

The F1 score, which incorporates the trade-off between precision and recall, also confirmed the superiority of ConvNeXt V2 Tiny Remod with a score of 98.1%. This balance shows the model’s general performance in the classification task; the model could classify most positive cases without classifying many as negative. On the same note, the F1 score of Swin Tiny at 69.9% indicates the model’s inability to balance between detecting true positives and minimising false positives ideally, thus being less suitable for the fine-tuning task of malaria detection.

The accuracy and precision of the ConvNeXt models are not the only factors that make them stand out. The ROC-AUC value of the model that quantifies how well the model can differentiate parasitised and uninfected samples was 0.996 for ConvNeXt V2 Tiny Remod. This high value points to the model being well-equipped to distinguish between the two categories. However, the ROC-AUC of 0.679 for Swin Tiny shows that it did not perform well in this regard, which reaffirms the previous observation that it was not quite adept at making the correct classifications.

In terms of confidence in the predictions of the model, the ConvNeXt V2 Tiny Remod was once more the best amongst the models, with a low log loss of 0.099. This low score suggests that the model made highly certain decisions, thus limiting the chances of misdiagnosis to the minimum. On the other end of the spectrum, with a log loss of 1.40, the Swin Tiny failed to distinguish between parasitised and uninfected image classes, thus playing into its subpar performance.

Additionally, ConvNeXt V2 Tiny Remod fared well in the MCC with a score of 0.962, which measures the overall performance of a model for both false positives and false negatives. This score enhances the credibility of the model in consistently producing accurate outcomes. Swin Tiny’s MCC was also relatively poor at 0.276, indicating that it was unreliable and had a high level of variability in classifying the samples. This was especially seen in the model’s specificity, which did not produce many false positive results. Based on the results, ConvNeXt V2 Tiny Remod had a specificity of 97.9%. Thus, it rarely offered false positive classifications of uninfected samples as parasitised, which is essential to avoid unnecessary treatments for patients who do not have the disease.

The sensitivity for Swin Tiny was 33.2%, a lower value compared to the other algorithms; this can be attributed to the model’s tendency to classify uninfected cells as parasitic, thus reducing the model’s usefulness in clinical diagnosis. Regarding balanced accuracy, that is, the mean of recall and specificity, ConvNeXt V2 Tiny Remod occupies the leading position with a score of 98.1%. This high value indicates the model’s good performance on both the positive and negative classes because it can distinguish between them and avoid over-prediction cases where it may erroneously predict the image as having parasites when it does not. This is because the balanced accuracy of Swin Tiny stands at a low 61.4%, which shows comprehensive inefficiency regarding balance.

The Cohen’s kappa scores of the ConvNeXt models also showed that the models made almost accurate classifications of the images. The ConvNeXt V2 Tiny Remod achieved an accuracy of 0.962, which shows a high correlation with the true labels; on the other hand, Swin Tiny, with an accuracy of 0.228, shows a low correlation and is in line with the earlier findings that it performed poorly.

Considering G-Mean, a measure combining recall and specificity, ConvNeXt V2 Tiny Remod is the best, with a value of 0.981. This implies that the model could distinguish between the parasitised images and, at the same time, minimise over-prediction. For instance, Swin Tiny, with a G-Mean of 0.545, failed to strike this balance, affecting its performance.

These findings indicate that the ConvNeXt models are substantially better than the others for detecting malaria parasites in microscopic images, especially the ConvNeXt V2 Tiny Remod model. Such models provided high accuracy and precision and had low error rates, thus showing indications of suitability for practical use. Conversely, Swin Tiny could have been stronger in several aspects, limiting its capability to address this classification problem.

The heatmap in Figure 7 supports the tabulated results as they compare model performance across multiple metrics. Of all the models, ConvNeXt V2 Tiny Remod and ConvNeXt Tiny have the best accuracy, precision, recall, and F1 score, as well as low log loss, thereby proving to be efficient in identifying parasitised and uninfected blood smears. This is supported by their high specificity and moderate sensitivity, which enhances their performance. Conversely, Swin Tiny and ConvNeXt V2 Tiny models show less stable results with lower precision and higher false positive rates, thus weaker classification capabilities. The general comparison indicates that ConvNeXt V2 Tiny Remod is the most viable model for malaria identification.

**Figure 7:**
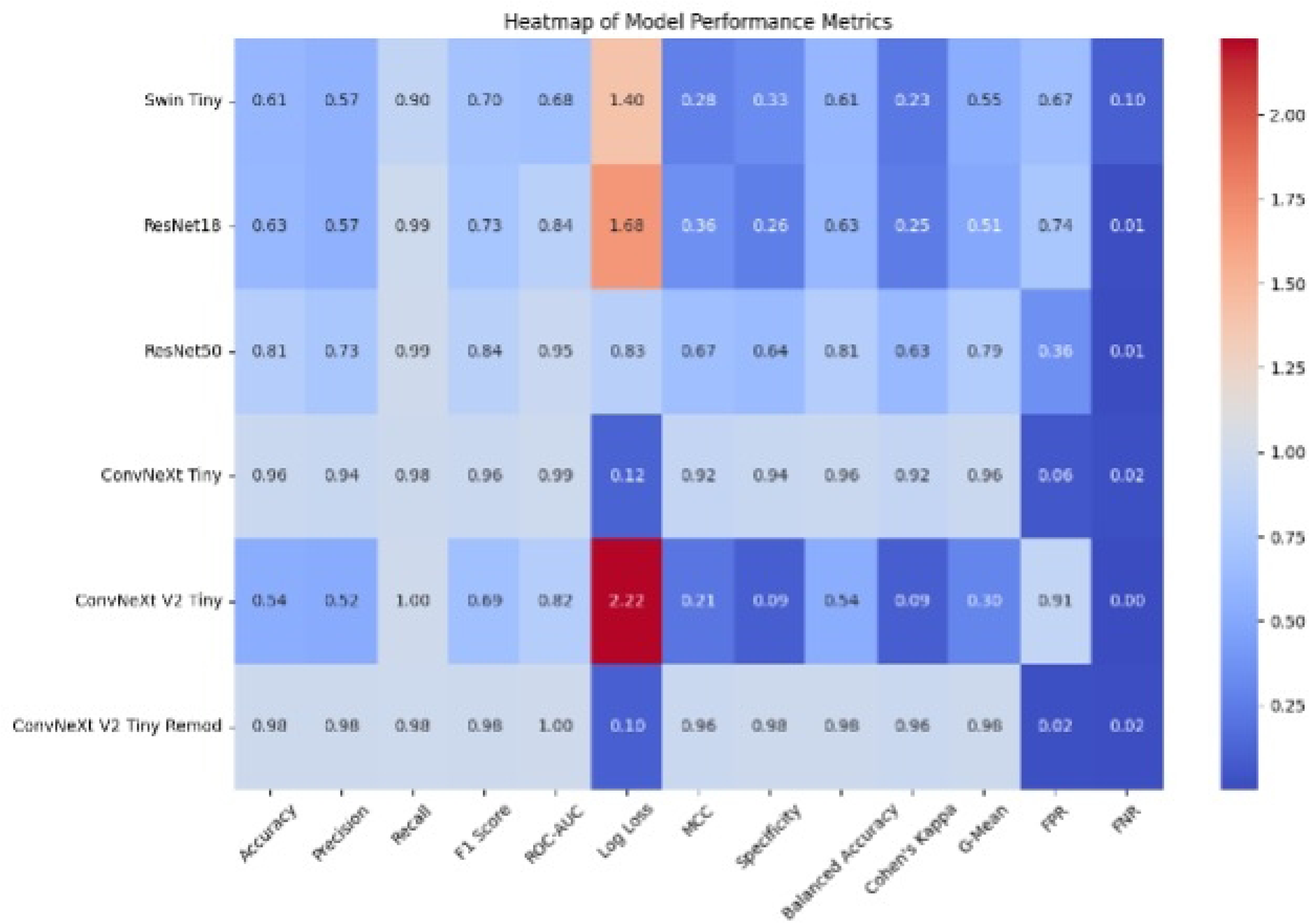
Heatmap of model performance metrics

This assessment further shows that the ConvNeXt models, especially the ConvNeXt V2 Tiny Remod, are more effective for malaria parasite detection in the blood smear slides. These models have high accuracy, precision, and balanced classification for medical imaging tasks and, therefore, have a great potential for enhancing diagnostic accuracy.

## 4. Discussion

This study shows that ConvNeXt architecture has many benefits and helps identify malaria parasites in thin blood smear images. The architectural design of ConvNeXt for high-resolution images allows it to have detailed local information and overall contextual information in the images. This two-fold function is beneficial in tasks involving basic and comprehensive information, such as medical diagnosis, since the images’ small details and overall picture must be analysed.

There are several reasons for the enhanced performance of ConvNeXt and, especially, the ConvNeXt V2 Tiny Remod model. First, the hierarchical feature extraction of the model can capture details of malaria parasites, including their shape, texture, and internal structure, while also capturing the general composition of the blood smear. This is very important in medical imaging, where diagnosis accuracy may depend on the finer details based on the image displayed.

Although this study’s dataset was from Bangladesh and thus restricts the geographic generalisability, data augmentation was applied to introduce variability. Future work should include datasets from other regions with different malaria strains to improve the model’s robustness. Collectively, the model could be expanded through global collaborations to improve its performance across different geographic contexts.

Another consideration for AI-based diagnostic systems is the possibility of bias arising from geographically or demographically limited training data. The dataset of this study mainly comprised blood smear images exclusively from Bangladesh. It raised issues with genetic diversity biases, such as ethnic differences in blood types and malaria strains. There is the potential that deploying the model in various regions or with different populations can affect its performance. Future studies should use more diverse datasets collected from different parts of the world, from different blood types, and different parasite strains to increase the generalisability of the model and ameliorate this bias.

Transfer learning was employed in the development of models in order to enhance their performance. The ImageNet dataset used to pre-train the models in this work provided them with a good starting point for general visual features such as edges, textures and shapes. This helped reduce the requirement for a large amount of annotated malaria data and improved the learning on the task-specific dataset.

To increase the model’s robustness to poor-quality images that are more likely to be encountered in low-resource settings, the model was trained and tested on images with noise, rotations, and varying brightness, using data augmentation techniques. Furthermore, using many data augmentation techniques increased the model’s generalisation capability. This enhanced the model’s stability and guaranteed that the model’s performance could be optimal in various practical situations.

However, these methods have some drawbacks which have been identified to affect their effectiveness. A limitation is that these models require access to computational resources, which may be scarce in some environments, particularly in resource-limited settings. Using architectures such as Swin-transformers and ConvNeXt can be computationally intensive, and this poses a challenge in practical applications where technology and infrastructure may be inadequate. On the other hand, access to these methods as an online service, where the computation can be deferred to reputable online platforms such as Amazon web services and Google Cloud, can be used as an alternative to enable access regardless of locally used computational resources (92–94).

While data augmentation and transfer learning could solve some problems, the problem of dataset representativeness still exists. The balanced dataset used in this study may not completely reflect the variability that can be potentially observed in real-world clinical settings, like staining technique, sample quality, or parasite life cycle stages. Additionally, the ConvNext models used in this study are the less advanced versions, ConvNext Tiny and ConvNext V2 Tiny models, due to the unavailability of computational resources. Future work can involve training more advanced models, such as ConvNext V2 XLarge, on more diverse and large datasets to find a way to generalise better.

The results from this study reveal the ability of ConvNeXt models, particularly in relation to the high recall and F1 scores that characterise these models and which are particularly useful for identifying malaria parasites. However, the utilisation of transfer learning and augmentation has drawbacks, including sensitivity to certain data distributions. This was somewhat offset by incorporating regularisation techniques such as the dropout.

Furthermore, including a new malaria diagnostic application within this framework emphasises the clinical significance of this study. The app builds upon ConvNeXt and incorporates methods such as LIME to facilitate real-time explainable AI diagnosis to healthcare professionals. This tool is especially useful in areas with a scarcity of specialists who are usually required to administer some of these tests. However, the app’s deployment might be restricted by the requirement for considerable computing ability and constant connectivity to the Internet, indicating more efficient field versions.

The ConvNeXt architecture shows clear clinical significance for malaria diagnosis in resource-limited environments. In high-burden areas, manual microscopy based on traditional methods is highly dependent upon skilled personnel and is prone to human error. An AI-based approach using ConvNeXt models to solve malaria detection has been proposed. It is automated and accurate in detecting malaria parasites and simultaneously tackles some of the key limitations of traditional approaches.

Additionally, the high accuracy and precision of ConvNeXt V2 Tiny Remod (98% accurate) demonstrate that deep learning-based systems are reliable for detecting malaria with accuracy similar to expert-level diagnosis. Such reliability may significantly alleviate the diagnostic workload of high-volume clinics and facilitate speeding up the diagnostic process, offering timely treatment and improving patient outcomes. Adding the layer of clinical significance to explainable AI techniques like LIME (for visual explanations) allows healthcare professionals to understand the AI model’s decision-making process. This puts into place an interpretability which builds trust with clinicians so that system outputs are both accurate and transparent.

## 5. Conclusion

Deep learning models, especially the ConvNeXt-based malaria detection models proposed in this study, are promising for real-world application, especially in low-resource settings. Malaria remains a major disease burden, especially in developing countries where there is a shortage of expert clinicians and well-equipped laboratories. The ConvNeXt V2 Tiny Remod model presented in this study with an accuracy of 98% presents an efficient method of automating malaria diagnosis. The scientific rationale is that ConvNeXt can be used to identify malaria parasites in blood smears because the architecture enables the model to capture detailed and contextual information as a microscopist would while examining blood smears under a microscope. This makes it very sensitive and specific in detection, which is vital in the early stages and correct identification of the parasites.

Moreover, data augmentation and transfer learning are implemented in the model, which improves the model’s performance regardless of the imaging conditions. These models could help decrease diagnostic errors, reduce the time to diagnosis, and enhance the clinical management of malaria patients. The use of diagnostic services in mobile or edge devices also presents a way of extending the services to regions without access to such services, thus contributing to efforts to fight this global disease burden.

## Data Availability

All image files used in this study are available from https://lhncbc.nlm.nih.gov/LHC-research/LHC-projects/image-processing/malaria-datasheet.html

